# Treatment with Arbidol and Moxifloxacin in Ordinary and Severe Adult Patients Infected with COVID-19

**DOI:** 10.1101/2020.05.30.20117598

**Authors:** Wen-Na Xi, Di Jin, Ke Sun, Rong-Yan Yu, Xue-Bing Yao, Biao-Shu Zou, Zhi-Ying Song, AO-Yu Yang, Rui-Xia Luo, Yun liu, Yan-hua Li, Shui-Lin Sun, Dong-Shan Yu

## Abstract

**Background:** An outbreak of coronavirus disease 2019 (COVID-19) caused by severe acute respiratory syndrome coronavirus 2 (SARS-CoV-2) has been widely spread. We aim to investigate the therapeutic effect of arbidol and moxifloxacin in patients infected with SARS-CoV-2.

**Methods:** We collected and analyzed data on 94 patients with COVID-19 including 27 severe patients at the Intensive Care Unit (ICU) and 74 ordinary patients at general isolation ward in Wuhan Xiehe Hospital, from February 15, 2020 to March 15, 2020. All patients were treated with arbidol (100mg each time, three times a day for 14 days) and moxifloxacin (0.4g each time, once a day for 7-14 days). Other data was also collected including demographic data, symptoms, laboratory findings, treatments and clinical outcomes.

**Results:** In basic characteristics, compared with the ordinary patients, the severe patients were older (median age was 63.0 years V.S 57.0 years, *p*=0.03), had higher proportion of hypertension (30% V.S 9%, *p*=0.03), higher possibility of getting fatigue and/or myalgia (26% V.S 6%, *p*=0.03), and had more obvious dyspnea symptom (26% V.S 3%, *p*=0.006). In regarding to laboratory results, we found the severe patients have higher white blood cell counts (p=0.003), neutrophil counts (p=0.007), higher levels of D-dimer (*p*<0.001), ALT (*p*<0.001) and AST (p=0.013) than the ordinary patients. After treatment of arbidol and moxifloxacin for one week, the rates of SARS-CoV-2 nucleic acid turning negative were 69.2% in the severe group and 77.8% in the ordinary group. A peculiar phenomenon was that IL-6 stands out among the cytokines in both groups, and higher in severe group than the ordinary one (p=0.011). After treating with arbidol and moxifloxacin for one week, IL-6 decreased significantly in severe group (p=0.023).

**Conclusion:** In summary, we proved the treatment of arbidol and moxifloxacin could be helpful in reducing viral load and inflammation during SARS-CoV2 infection, especially for negatively regulating fatal inflammation in severe COVID-19 patients. However, more evidence awaits further clinical verification.

## INTRODUCTION

A novel coronavirus designated as SARS-CoV-2 has been known causing an international outbreak of respiratory illness named COVID-19 (Huang et al., 2020; Wang et al., 2020). Up to May 31, 2020, more than 5.75 million people in worldwide has been confirmed with COVID-19, at least 361 thousand patients died (Johns Hopkins University - https://coronavirus.jhu.edu/map.html). Effective and specific treatment for COVID-19 is the most urgent task at the moment. Up to now, there is still no specific treatment for SARS-CoV-2 infection. Several drugs, such as chloroquine, hydroxychloroquine, lopinavir/ritonavir, have shown some effects in treating COVID-19 (Huang et al., 2020; Meo et al., 2020; Wang et al., 2020), however also come along with serious side effects.

Arbidol is a synthetic broad-spectrum antiviral agent that is used to treat many virus infections in both clinic and experimental research, such as seasonal influenza, ebola virus, SARS-CoV, lassa viruses and paramyxo (Shi et al., 2007; Blaising et al., 2013; Blaising et al., 2014b; Pecheur et al., 2016). So far, little is known about application of abidor in the treatment of COVID-19. Moxifloxacin is a fourth-generation fluoroquinolone with expanded active against a wide range of aerobic gram-positive and gram-negative organisms, which is commonly used and suitable for respiratory bacterial infection (Ding and Zhang, 2018).

To date, clinical evidence on arbidol and moxifloxacin in patients with COVID-19 is limited. Herein, we evaluated the therapeutic effects of arbidol and moxifloxacin in patients with COVID-19.

## MATERIAL AND METHODS

### Patients

We included 67 ordinary patients at general isolation ward and 27 severe patients at the Intensive Care Unit (ICU) from Wuhan Xiehe Hospital (admission data from February 15 to March 15). COVID-19 was clinically diagnosed as “viral pneumonia” based on clinical symptoms, epidemiological exposure history, chest computed tomographic (CT) scan and real-time reverse transcription-polymerase chain reaction (RT-PCR) assay for SARS-CoV-2 from throat swab specimens. Clinical stage of the patients was classified according to “Interim Guidance for Diagnosis and Treatment of Coronavirus Disease 2019 (the 6th edition)” released by National Health Commission.

Severe COVID-19 was defined when patients got one of the following criteria: (1) respiratory frequency ≥ 30 times/min; (2) oxygen saturation < 93% at rest; (3) arterial partial pressure of oxygen (PaO2)/oxygen concentration (FiO2) ≤ 300 mmHg (1 mmHg = 0.133 kPa).

### Laboratory testing

SARS-CoV-2 viral nucleic acid was detected by RT-PCR assay from pharyngeal swab specimens of the patients at Wuhan Xiehe hospital. The detailed protocol was described before (Huang et al., 2020). IgM and IgG antibodies against SARS-CoV-2 were measured by SARS-CoV-2 IgG/IgM antibody test kit (C86095G/C86095M, YHLO biotech, Shenzhen) according to the protocol. Medical laboratory data was collected, including a complete blood count (leucocytes, neutrophils, lymphocytes), serum biochemistry results, such as levels of alanine aminotransferase (ALT), aspartate aminotransferase (AST), D-dimer, and inflammatory cytokines (IL-2, IL-4, IL-6, IL-10, TNF-α and IFN-γ). All medical laboratory data were generated from the Department of Clinical Laboratory of Wuhan Xiehe Hospital.

### Statistical analysis

Categorical data was summarized as frequencies (%) and compared by χ^2^ test or Fisher’s exact test between ordinary and severe groups. Continuous variables were described as median and interquartile ranges (IQR) values and compared with the Mann-Whitney U test. All analyses were performed with Graphpad Prism 5 (GraphPad Software, UK). Differences with p < 0.05 between group means were considered statistically significant.

## RESULTS

### Demographic and baseline characteristics

Our study involved 94 cases of COVID-19 patients, including 67 ordinary and 27 severe patients. As shown in table 1, the median age of severe cases was younger than the ordinary cases (P=0.03). In comorbidity, hypertension was more common in the severe group (P=0.03). Meanwhile, for clinical symptoms, larger proportion of patients had fatigue or myalgia, and dyspnea in severe group (P=0.006). Fever (50%) and cough (31%) are the most common symptoms at onset of the illness. There was not enough evidence in this study showed that there were statistical differences in numbers of chronic pulmonary disease, cardiovascular disease and diabetes in each group.

**Table 1.** Demographics and baseline characteristics of patients infected with COVID-19.

| Basic characteristics | All patients<br>(n=94) | Ordinary cases<br>(n=67) | Severe cases<br>(n=27) | P value |
| --- | --- | --- | --- | --- |
| Age, years | 61 (49, 61) | 57 (45, 65) | 63 (57, 76) | 0.03 |
| Sex |  |  |  |  |
| Women | 58 (62%) | 46 (63%) | 12 (44%) | - |
| Men | 36 (38%) | 21 (37%) | 15 (56%) | - |
| Exposure history |  |  |  |  |
| family cluster | 3 (3%) | 2 (3%) | 1 (4%) | 0.87 |
| Any comorbidity |  |  |  |  |
| Chronic pulmonary disease | 7 (7%) | 4 (6%) | 3 (11%) | 0.39 |
| Cardiovascular disease | 5 (5%) | 1 (2%) | 4 (15%) | 0.07 |
| Hypertension | 14 (15%) | 6 (9%) | 8 (30%) | 0.03 |
| Chronic liver disease | 6 (6%) | 3 (4%) | 3 (11%) | 0.24 |
| Diabetes | 7 (7%) | 5 (8%) | 2 (7%) | 0.99 |
| Signs and symptoms |  |  |  |  |
| Fever | 47 (50%) | 34 (51%) | 13 (48%) | 0.51 |
| Highest temperature, °C |  |  |  |  |
| 37.3–38.0 | 28 (30%) | 20 (30%) | 8 (30%) | - |
| 38.1–39.0 | 13 (14%) | 10 (15%) | 3 (11%) | - |
| >39.0 | 6 (9%) | 4 (6%) | 2 (7%) | - |
| Cough | 29 (31%) | 18 (27%) | 11 (41%) | 0.22 |
| Fatigue or Myalgia | 11 (12%) | 4 (6%) | 7 (26%) | 0.03 |
| Dyspnea | 9 (10%) | 2 (3%) | 7 (26%) | 0.006 |
| Diarrhoea | 3 (3%) | 2 (3%) | 1 (4%) | 0.87 |
| Note |  |  |  |  |

### Clinical characteristics

As table 2 shown, the blood counts of patients on admission showed an uncommon phenomenon that white blood cell (WBC) counts in severe patients (median, 7.2×10^9^/L) were much higher than ordinary patients (median, 5.1×10^9^/L) (P=0.003). D-dimer level on admission were significantly higher in severe patients (median, 3.2 mg/L) than ordinary patients (median, 0.8 mg/L) (*P*< 0.001). Meanwhile, severe cases induced more significant liver damage (ALT median, 108.4 U/L, AST median, 74.6 U/L) than ordinary cases (ALT median, 42.4 U/L, AST median, 36.3 U/L) (*P*< 0.001, =0.013, respectively).

**Table 2.** Laboratory findings of patients infected with COVID-19.

| Laboratory tests | All patients<br>(n=94) | Mild cases<br>(n=67) | Severe cases<br>(n=27) | P value |
| --- | --- | --- | --- | --- |
| White blood cell count,<br>× 10<sup>9</sup>/L | 5.7 (4.1-6.3) | 5.1(3.9-6.3) | 7.2 (4.6-7.9) | 0.003 |
| <4 | 11 (12%) | 8 (12%) | 3 (11%) | - |
| 4-10 | 80 (85%) | 59 (88%) | 21 (78%) | - |
| >10 | 3 (3%) | 0 | 3 (11%) | - |
| Neutrophil count, × 10<sup>9</sup>/L | 3.8 (2.3-4.2) | 3.2 (2.2-3.7) | 5.1 (2.6-5.4) | 0.007 |
| Lymphocyte count, × 10<sup>9</sup>/L | 1.3 (0.8-1.6) | 1.3 (0.9-1.7) | 1.1 (0.8-1.3) | 0.113 |
| <1.0 | 23 (24%) | 11 (16%) | 12 (44%) | - |
| ≥1.0 | 71 (76%) | 56 (84%) | 15 (56%) | - |
| D-dimer, mg/L | 1.5 (0.3-1.7) | 0.8 (0.3-1.0) | 3.2 (0.5-5.3) | < 0.001 |
| Alanine aminotransferase<br>(ALT), U/L | 61.4 (33.0-62.0) | 42.4 (32.0-47.0) | 108.4 (45.0-142.0) | < 0.001 |
| Aspartate aminotransferase<br>(AST), U/L | 47.2 (23.0-55.0) | 36.3 (23.0-44.0) | 74.6 (33.0-78.0) | 0.013 |
| T-lymphocyte subsets<br>(on admission) |  |  |  |  |
| CD3+ (%) | 72.1 (66.3-80.2) | 72.1 (66.2-82.4) | 75.1 (66.2-80.2) | 0.96 |
| CD4+ (%) | 49.6 (42.1-55.3) | 48.6 (41.5-55.4) | 51.5 (45.8-54.3) | 0.285 |
| CD8+ (%) | 24.6 (20.0-29.1) | 25.5 (21.1-27.3) | 23.0 (15.6-30.2) | 0.214 |
| CD4+/CD8+ | 2.3 (1.7-2.6) | 2.0 (1.6-2.6) | 2.9 (1.8-3.3) | 0.014 |
| Positive COVID-19 nucleic<br>acid |  |  |  |  |
| Admission | 88 (94 %) | 61 (91%) | 27 (100%) | - |
| 7 days after admission | 40 (43%) | 27 (40%) | 13 (48%) | - |
| 14 days after admission | 10 (11%) | 6 (9%) | 4 (15%) | - |
| COVID-19 IgM, positive | 45 (48%) | 33 (49%) | 12 (44%) | - |
| COVID-19 IgG, positive | 31 (33%) | 26 (39%) | 5 (19%) | - |
| Note: |  |  |  |  |

### Treatment with arbidol and moxifloxacin

We choose arbidol and moxifloxacin as the therapy. In ordinary group, 61 cases (91%) got positive SARS-CoV-2 viral nucleic acid when at admission. Surprisingly, 27 cases (40%) remained positive after one-week treatment and only 6 cases (9%) kept positive two weeks later. In severe group, 27 cases (100%) got positive SARS-CoV-2 viral nucleic acid at admission, 13 cases (48%) remained positive one week later and 4 cases (15%) positive two week later. Although we did not set a placebo group, the treatment was still obviously effective. It is worth mentioning that 33 cases (49%) in ordinary group and 12 cases (44%) in severe group got positive SARS-CoV-2 IgM at admission, while 26 cases (39%) got positive IgG in ordinary group and 5 (19%) cases got positive SARS-CoV-2 IgG in severe group at admission (table 1). Unfortunately, we were unable to follow-up with the IgM and IgG data later due to non-science reasons.

Meanwhile, we checked six common cytokines, showed in table 3, including IL-2, IL-4, IL-6, IL-10, TNF-α, IFN-γ. Interestingly, IL-6 increased significantly in severe group compared to ordinary group (P=0.011), while IL-2, IL-4, IL-10, TNF-α, IFN-γ were all at a low level in both ordinary and severe patient’s groups (figure 1). However, in severe group, IL-6 significantly decreased after one-week treatment with arbidol and moxifloxacin (P=0.023) (figure 2).

**Table 3.**
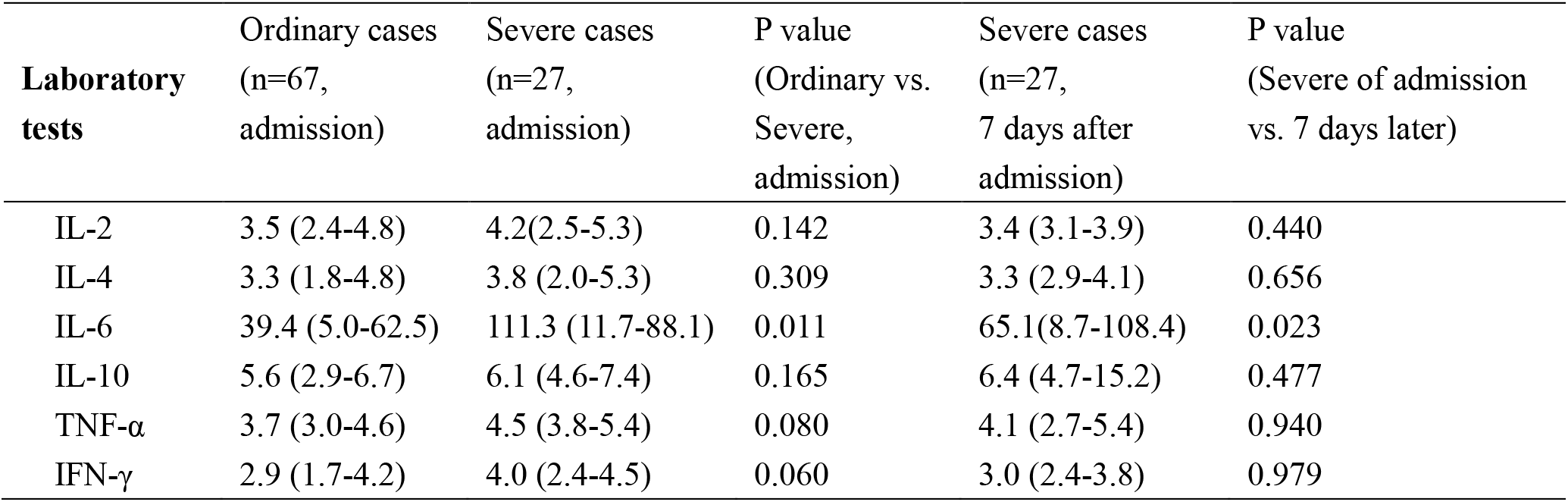
Plasma cytokines assay of patients infected with COVID-19.

**Figure 1.**
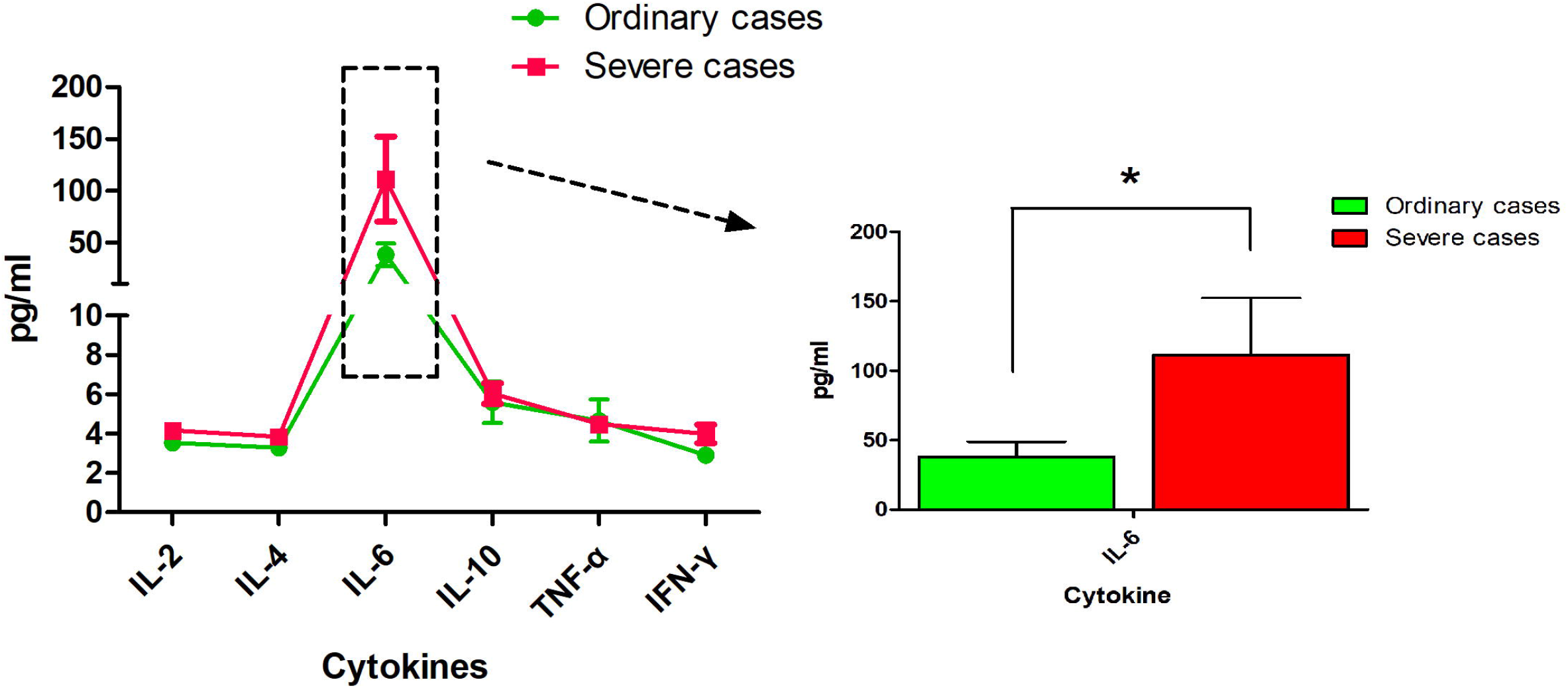
The levers of six cytokines (IL-2, IL-4, IL-6, IL-10, TNF-α and IFN-γ) in ordinary and severe groups of COVID-19 at admission. IL-6 got significantly higher in severe cases compared to ordinary cases (P=0.011), but IL-2, IL-4, IL-10, TNF-α and IFN-γ were all at a low level in both ordinary and severe groups.

**Figure 2.**
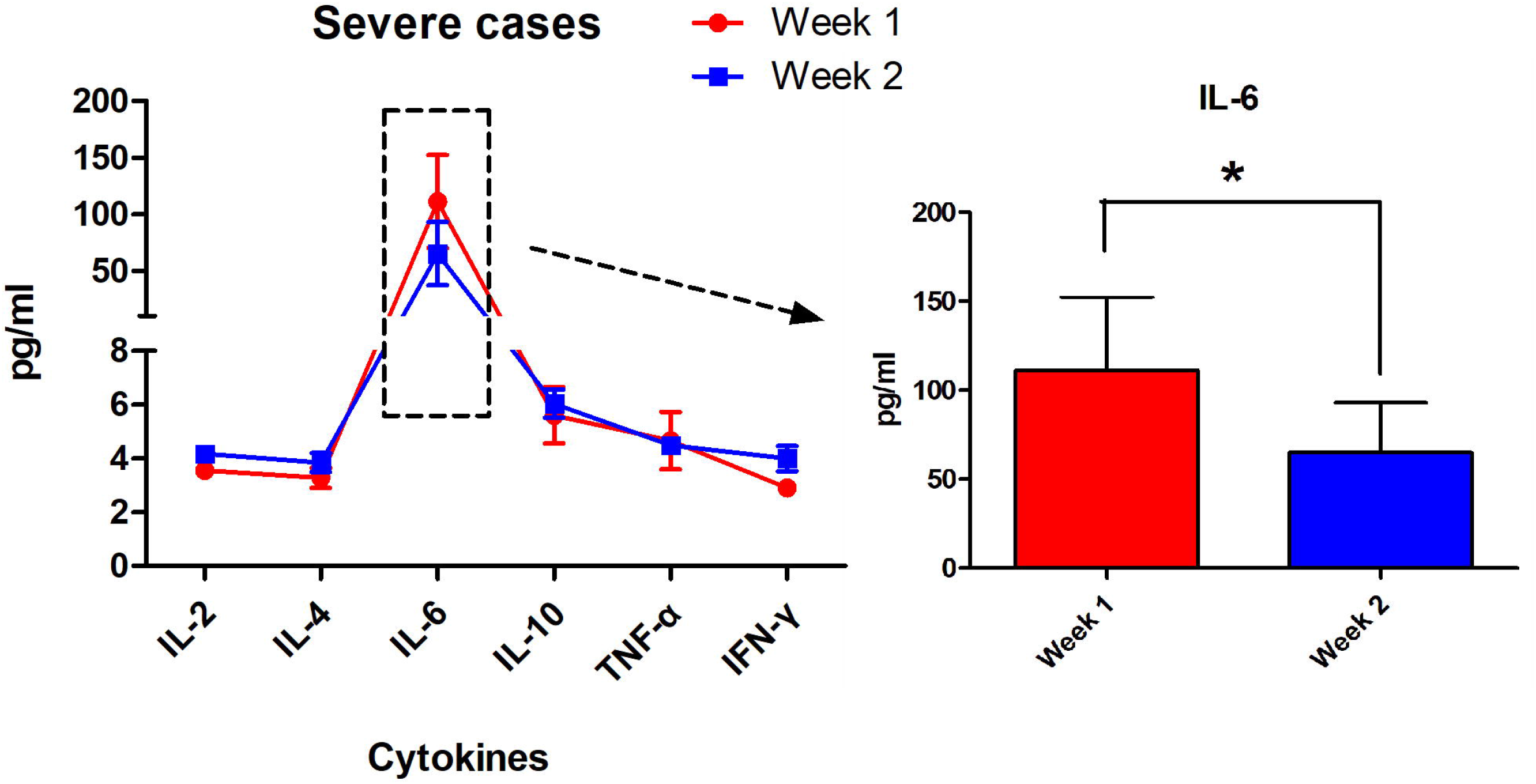
Changes in levels of six cytokines before and after treatment for one-week. In severe group, IL-6 significantly decreased after one-week treatment with arbidol and moxifloxacin (P=0.023), while IL-2, IL-4, IL-10, TNF-α and IFN-γ got little changes.

## DISCUSSION

In the present study, we provided clinical data of 94 COVID-19 infections, including both primary cases and severe cases. Similar to other studies, our data indicated senior and hypertension are the high risk factors for severe COVID-19 infection. Meanwhile, fatigue, myalgia and dyspnea were common symptoms in patients of severe cases.

In clinical features, lymphopenia was common in patients of both groups (75.4%). High level of D-dimer, ALT and AST in severe group indicating sustained inflammatory response and disturbed coagulation mechanism after infection with SARS-CoV-2 and this phenomenon was severer in fatal cases.

Cytokines storm is one of the characteristics of serious infectious disease, especially some viral infectious diseases, such as highly pathogenic avian influenza H5N1(Huo et al., 2018). As well known that IFN-α/-β, TNF-α, IL-1, IL-6˴ IL-8˴ IL-10 and other cytokines account for the major proportion in the storm and trigger violent inflammatory response, CD3+, CD4+, and CD8+ T cells significantly change at the same time (Chi et al., 2013; Liu et al., 2016; Guo and Thomas, 2017). In our cohort, we were surprised that IL-2, IL-4, IL-10, TNF-α, IFN-γ were all at a low or even normal levels both in ordinary and severe COVID-19 patients, but IL-6 increased sharply and significantly higher in severe group than ordinary group. This had some discrepancies with reports in other similar literatures (Sun et al., 2020; Zhao, 2020). Meanwhile, the count of CD3+, CD4+, and CD8+ T cells in the cohort, both in ordinary and severe group, were almost always within the normal range.

It is well known that IL-6 is normally secreted by Mononuclear/macrophages, T lymphocytes, B lymphocytes and epithelial cells. When infection and inflammation occurs, IL-6 generation goes first and increases rapidly, which is consistent with the severity of the infection and can be used as a sensitive indicator for the early diagnosis of acute infection (Puel and Casanova, 2019; Liu, 2020). This may explicit there was higher level of IL-6 in severe COVID-19 patients. However, it is difficult to understand that important inflammatory cytokines including IL-10, TNF-α and IFN-γ got little response in the course. Does immune escape involve in COVID-19 virus infection. Further and larger scale clinical research is drastically needed in deed. Meanwhile, in severe group, the level of IL-6 seriously decreased after one-week treatment with arbidol and moxifloxacin (P=0.023), which indicated this therapy effectively reduced the inflammatory level of host. This was consistent with what already have been known about Arbidol. Arbidol is an indole-based molecule able to interactions with selective amino-acid residues of proteins (phenylalanine, tyrosine, tryptophan), impair several steps in the life cycle of viruses including attachment to cells, fusion of viral and cellular membranes (Teissier et al., 2011; Blaising et al., 2014a). Arbidol has been proved to inhibit influenza virus, HCV, ZIKV and EBOV(Boriskin et al., 2006; Kadam and Wilson, 2017; Fink and Vojtech, 2018; Hulseberg et al., 2019). Based on previous information and our results, we hypothesized that arbidol could inhibit the SARS-CoV-2 virus infection, however more relevant basic research is still seriously needed.

Viral pneumonia is usually accompanied by bacterial infection (Babiuk et al., 1988; Zhou et al., 2018), and COVID-19 is not an exception. Prevention and treatment of bacterial infection benefits the recovery from viral pneumonia. Moxifloxacin has strong antibacterial properties, broad antibacterial spectrum and few adverse reactions, makes it suitable for the treatment of lung infection (Rijnders, 2003; Öbrink-Hansen et al., 2015). In our cohort, we combined arbidol and moxifloxacin to treat COVID-19 patients. 51% of the patients in ordinary group and 52% in severe group became negative in viral nucleic acid tests, after one week’s treatment, surprisingly, 82% of the patients in ordinary group and 85% in severe group turned negative in viral nucleic acid tests after 2 weeks of treatment. Meanwhile, IL-6 decreased significantly in severe group after one week’s treatment. Taken together, these data indicated the positive effects of arbidol and moxifloxacin on COVID-19 treatment.

In summary, the study accumulated clinical and laboratory characteristics of 94 COVID-19 patients, especially changes in SARS-CoV-2 nucleic acid and inflammatory cytokines after treatment of arbidol and moxifloxacin. And we proved the positive efficacy on COVID-19 with treatment of arbidol and moxifloxacin. However, as the size of this cohort is limited, the statistical analysis results should be interpreted with caution and need further experimental and clinical verification.

## Data Availability

The authors confirm that the data supporting the findings of this study are available within the article.

## ACKNOWLEDG MENTS

We greatly acknowledge all the healthcare professionals who took care of the COVID-19 patients with their great effort, especially WN Xi and K Sun, who went to Wuhan Xiehe hospital to support the fight against the COVID-19 epidemic. Meanwhile, this work was supported by grants from the National Science and Technology Major Project for the Control and Prevention of Major Infectious Diseases in China (201811300954127).

## DECLARTION OF INTERESTS

We declare no competing interests.

## References

Babiuk L.A., Lawman M.J., and Ohmann H.B. (1988). Viral-bacterial synergistic interaction in respiratory disease. Adv Virus Res 35, 219–249. doi: 10.1016/s0065-3527(08)60713-7.

Blaising J., Levy P.L., Polyak S.J., Stanifer M., Boulant S., and Pecheur E.I. (2013). Arbidol inhibits viral entry by interfering with clathrin-dependent trafficking. Antiviral Res 100(1), 215–219. doi: 10.1016/j.antiviral.2013.08.008.

Blaising J., Polyak S.J., and Pécheur E.I. (2014a). Arbidol as a broad-spectrum antiviral: an update. Antiviral Res 107, 84–94. doi: 10.1016/j.antiviral.2014.04.006.

Blaising J., Polyak S.J., and Pecheur E.I. (2014b). Arbidol as a broad-spectrum antiviral: an update. Antiviral Res 107, 84–94. doi: 10.1016/j.antiviral.2014.04.006.

Boriskin Y.S., Pécheur E.I., and Polyak S.J. (2006). Arbidol: a broad-spectrum antiviral that inhibits acute and chronic HCV infection. Virol J 3, 56. doi: 10.1186/1743-422x-3-56.

Chi Y., Zhu Y., Wen T., Cui L., Ge Y., Jiao Y., et al. (2013). Cytokine and chemokine levels in patients infected with the novel avian influenza A (H7N9) virus in China. J Infect Dis 208(12), 1962–1967. doi: 10.1093/infdis/jit440.

Ding R.D., and Zhang H.J. (2018). Effect of linezolid on serum PCT, ESR, and CRP in patients with pulmonary tuberculosis and pneumonia. Medicine (Baltimore) 97(37), e12177. doi: 10.1097/md.0000000000012177.

Fink S.L., and Vojtech L. (2018). The Antiviral Drug Arbidol Inhibits Zika Virus. 8(1), 8989. doi: 10.1038/s41598-018-27224-4.

Guo X.J., and Thomas P.G. (2017). New fronts emerge in the influenza cytokine storm. 39(5), 541–550. doi: 10.1007/s00281-017-0636-y.

Huang C., Wang Y., Li X., Ren L., Zhao J., Hu Y., et al. (2020). Clinical features of patients infected with 2019 novel coronavirus in Wuhan, China. Lancet 395(10223), 497–506. doi: 10.1016/s0140-6736(20)30183-5.

Hulseberg C.E., Fénéant L., Szymańska-de Wijs K.M., Kessler N.P., Nelson E.A., Shoemaker C.J., et al. (2019). Arbidol and Other Low-Molecular-Weight Drugs That Inhibit Lassa and Ebola Viruses. 93(8). doi: 10.1128/jvi.02185-18.

Huo C., Xiao K., Zhang S., Tang Y., Wang M., Qi P., et al. (2018). H5N1 Influenza a Virus Replicates Productively in Pancreatic Cells and Induces Apoptosis and Pro-Inflammatory Cytokine Response. Front Cell Infect Microbiol 8, 386. doi: 10.3389/fcimb.2018.00386.

Kadam R.U., and Wilson I.A. (2017). Structural basis of influenza virus fusion inhibition by the antiviral drug Arbidol. Proc Natl Acad Sci USA 114(2), 206–214. doi: 10.1073/pnas.1617020114.

Liu A.Y. (2020). Infectious Implications of Interleukin-1, Interleukin-6, and T Helper Type 2 Inhibition. Infect Dis Clin North Am. doi: 10.1016/j.idc.2020.02.003.

Liu Q., Zhou Y.H., and Yang Z.Q. (2016). The cytokine storm of severe influenza and development of immunomodulatory therapy. Cell Mol Immunol 13(1), 3–10. doi: 10.1038/cmi.2015.74.

Meo S.A., Klonoff D.C., and Akram J. (2020). Efficacy of chloroquine and hydroxychloroquine in the treatment of COVID-19. Eur Rev Med Pharmacol Sci 24(8), 4539–4547. doi: 10.26355/eurrev_202004_21038.

Öbrink-Hansen K., Hardlei T.F., Brock B., Jensen-Fangel S., Kragh Thomsen M., Petersen E., et al. (2015). Moxifloxacin pharmacokinetic profile and efficacy evaluation in empiric treatment of community-acquired pneumonia. Antimicrob Agents Chemother 59(4), 2398–2404. doi: 10.1128/aac.04659-14.

Pecheur E.I., Borisevich V., Halfmann P., Morrey J.D., Smee D.F., Prichard M., et al. (2016). The Synthetic Antiviral Drug Arbidol Inhibits Globally Prevalent Pathogenic Viruses. J Virol 90(6), 3086–3092. doi: 10.1128/jvi.02077-15.

Puel A., and Casanova J.L. (2019). The nature of human IL-6. 216(9), 1969–1971. doi: 10.1084/jem.20191002.

Rijnders B.J. (2003). Moxifloxacin for community-acquired pneumonia. Antimicrob Agents Chemother 47(1), 444; author reply 444–445. doi: 10.1128/aac.47.1.444-445.2003.

Shi L., Xiong H., He J., Deng H., Li Q., Zhong Q., et al. (2007). Antiviral activity of arbidol against influenza A virus, respiratory syncytial virus, rhinovirus, coxsackie virus and adenovirus in vitro and in vivo. Arch Virol 152(8), 1447–1455. doi: 10.1007/s00705-007-0974-5.

Sun X., Wang T., Cai D., Hu Z., Chen J., Liao H., et al. (2020). Cytokine storm intervention in the early stages of COVID-19 pneumonia. Cytokine Growth Factor Rev. doi: 10.1016/j.cytogfr.2020.04.002.

Teissier E., Zandomeneghi G., Loquet A., Lavillette D., Lavergne J.P., Montserret R., et al. (2011). Mechanism of inhibition of enveloped virus membrane fusion by the antiviral drug arbidol. PLoS One 6(1), e15874. doi: 10.1371/journal.pone.0015874.

Wang D., Hu B., Hu C., Zhu F., Liu X., Zhang J., et al. (2020). Clinical Characteristics of 138 Hospitalized Patients With 2019 Novel Coronavirus-Infected Pneumonia in Wuhan, China. Jama. doi: 10.1001/jama.2020.1585.

Zhao M. (2020). Cytokine storm and immunomodulatory therapy in COVID-19: Role of chloroquine and anti-IL-6 monoclonal antibodies. Int J Antimicrob Agents, 105982. doi: 10.1016/j.ijantimicag.2020.105982.

Zhou S., Ren X., Yang J., and Jin Q. (2018). Evaluating the Value of Defensins for Diagnosing Secondary Bacterial Infections in Influenza-Infected Patients. Front Microbiol 9, 2762. doi: 10.3389/fmicb.2018.02762.

